# A clustering of missense variants in the crucial chromatin modifier WDR5 defines a new neurodevelopmental disorder

**DOI:** 10.1101/2021.11.01.21265518

**Authors:** Lot Snijders Blok, Jolijn Verseput, Dmitrijs Rots, Hanka Venselaar, A. Micheil Innes, Connie Stumpel, Katrin Ounap, Karit Reinson, Eleanor G. Seaby, Shane McKee, Barbara Burton, Katherine Kim, Johanna M. van Hagen, Quinten Waisfisz, Pascal Joset, Katharina Steindl, Anita Rauch, Dong Li, Elaine Zackai, Sarah Sheppard, Beth Keena, Hakon Hakonarson, Andreas Roos, Nicolai Kohlschmidt, Anna Cereda, Maria Iascone, Erika Rebessi, Kristin D. Kernohan, Philippe M. Campeau, Francisca Millan, Jesse A. Taylor, Hanns Lochmüller, Martin R. Higgs, Amalia Goula, Birgitta Bernhard, Simon E. Fisher, Han G. Brunner, Tjitske Kleefstra

## Abstract

WDR5 is a broadly studied, highly conserved protein involved in a wide array of biological functions. Among these functions, WDR5 is a part of several protein complexes that affect gene regulation via post-translational modification of histones. Here, we present data from ten unrelated individuals with six different rare *de novo* missense variants in *WDR5*; one identical variant was found in four individuals, and another variant in two individuals. All ten individuals had neurodevelopmental disorders including speech/language delays (N=10), intellectual disability (N=8), epilepsy (N=6) and autism spectrum disorder (N=4). Additional phenotypic features included abnormal growth parameters (N=6), heart anomalies (N=2) and hearing loss (N=2). All six missense variants occurred in regions of the *WDR5* locus that are known to be extremely intolerant for variation. Three-dimensional structures indicate that all the residues affected by these variants are located at the surface of one side of the WDR5 protein. It is predicted that five out of the six amino-acid substitutions disrupt interactions of WDR5 with RbBP5 and/or KMT2A/C, as part of the COMPASS family complexes. Thus, we define a new neurodevelopmental disorder associated with missense variants in *WDR5* and a broad range of associated features including intellectual disability, speech/language impairments, epilepsy and autism spectrum disorders. This finding highlights the important role of COMPASS family proteins in neurodevelopmental disorders.

## Introduction

WDR5 is a small highly conserved protein that is able to interact with a large number of other proteins^1,2^. As a core constituent of many different chromatin-related protein complexes, it controls crucial regulatory processes during development^3,4^. The indispensable function of WDR5 is illustrated by its high evolutionary conservation. Even very basic multicellular organisms such as *Trichoplax adhaerens* have a protein with around 90% similarity to the 334 amino acids of the human orthologue^4,5^. WDR5 is a member of the WD40 repeat family and has seven WD40 domains that each forms a propeller-like wing, resulting in a final barrel-shaped protein^6^. Using two binding sites on opposite sides of the protein, WDR5 can act as an adapter to link different proteins and form protein complexes. Since the protein is highly multifunctional and ubiquitously expressed^7^ (data available from https://v19.proteinatlas.org/ENSG00000196363-WDR5), the unavailability of a well-functioning WDR5 could potentially impact myriad downstream processes.

Most of the protein complexes that WDR5 participates in affect gene regulation via post-translational modification of histones. The MLL/SET complexes (also known as COMPASS family complexes) catalyse histone 3 lysine 4 (H3K4) di- and trimethylation^8,9^ and the NSL and ATAC complexes are involved in histone acetylation^10,11^. WDR5 can also be part of an embryonic stem cell-specific NuRD complex that combines nucleosome sliding capacities with histone deacetylation activity^12^. In addition to interactions with other proteins, WDR5 is also able to bind to >1000 different endogenous RNAs (including long non-coding RNAs), and binding to certain long non-coding RNAs can be crucial for WDR5 stability and function in cells^13^. A recent study linked WDR5, as part of the COMPASS complex, to a newly discovered genetic compensation mechanism: nonsense-induced transcriptional compensation^14^. In short, this mechanism is triggered by a truncating variant in a gene, and leads to the expression of related genes, thereby compensating for the effects of a deleterious variant^15^. One of the most well-studied aspects of WDR5 function is its role in embryonic stem cell (ESC) self-renewal and maintenance of a pluripotent state^16,17^. Recently, a direct interaction between the p53 protein and WDR5 has been uncovered, in which mouse ESC stem cell fate (the differentiation into neuroectoderm or mesoderm) is regulated in a p53-dependent manner^18^. Thus, WDR5 has already been implicated in multiple different molecular pathways and mechanisms, and this list is growing steadily.

While the biological functions of the WDR5 protein have been studied from numerous angles, there is still little known about the impact of rare germline variants in the gene that encodes it. Using data from large-scale sequencing resources, it is clear that *WDR5* is extremely intolerant for loss-of-function variation: in both the gnomAD database^19^ (version 2.1.1; containing sequencing data of 141,456 individuals) and the TOPMED database^20^ (containing sequencing data of 62,784 individuals) not a single truncating variant in *WDR5* is reported. Similarly, *WDR5* is also depleted for missense variants. Therefore, the initial finding of a *de novo* missense variant (p.(Thr208Met)) in *WDR5* in a child with a developmental speech disorder^21^, prompted us to investigate the effects and possible pathogenicity of rare germline variants in this gene. We collected clinical information on this proband and nine additional individuals with rare *de novo* germline variants in *WDR5*, collated from several clinical exome or genome sequencing studies. We used a range of *in silico* tools and analyses of variants using three-dimensional structures in order to evaluate the likely consequences of the different variants found.

## Methods

### Study participants and consent

Individuals with *WDR5* variants were identified via matchmaking using GeneMatcher^22^, the Dutch genetic diagnostic variant classification database (VKGL database)^23-25^, ClinVar^26^ and denovo-db^27^. Clinical data and details on variants were collected in a Castor EDC database^28^. Informed consent to share and publish these data was given by all individuals or their legal guardian. Consent for publication of photographs was obtained separately. Genetic testing and research were performed in accordance with protocols approved by the local Institutional Review Boards.

### Next generation sequencing and *in silico* variant analyses

Details on next generation sequencing methods used to identify the *WDR5* variants found in all individuals are included in table S1. Variants were analysed using Alamut Visual 2.10. Conservation was studied using a Clustal^29^ alignment of WDR5 amino acid sequences extracted from Uniprot (human, mouse and C. elegans)^30^. To assess the likelihood of pathogenicity, the prediction programs SIFT^31^, PolyPhen-2^32^ and CADD v1.4^33^ were used.

### Three-dimensional (3D) protein modelling

The effects of the identified variants on the WDR5 protein and its interaction with other proteins in the COMPASS family complexes were analyzed using YASARA View^34^ with FoldX v4.0 plugin^35^. For the WDR5 structure, PDB file 2GNQ was used. PDB files 6KIV and 6KIW^36^ were used for the analysis of the core MLL1 and MLL3 complexes, respectively; the 6UH5 file^37^ of the yeast COMPASS model was used for the comparison with the human MLL1 complex. To optimize the position of amino-acid sidechains, all the PDB files that were used were corrected by the FoldX repair function using default settings. Different protein structures were aligned with SHEBA procedure^38^, as implemented in YASARA.

## Results

### Identification and clinical characterization of individuals with *de novo WDR5* variants

We report clinical and molecular data on ten unrelated individuals with a *de novo* missense variant in *WDR5*. Six different missense variants were reported in these ten individuals: p.(Ala169Pro), p.(Arg196Cys), p.(Ala201Val), p.(Thr208Met), p.(Asp213Asn) and p.(Lys245Arg). The p.(Thr208Met) variant was found in four unrelated individuals, and the p.(Arg196Cys) variant in two unrelated individuals.

All individuals had neurodevelopmental disorders with a spectrum of overlapping associated features (Figure 1 and table S1). Intellectual disability (ID) was present in eight out of ten individuals, with a severity ranging from moderate ID (IQ 35-50, five individuals) to mild ID (IQ 50-70, three individuals). Of the two remaining individuals, one individual had a borderline level of intellectual functioning (IQ 70-85) and another individual had no intellectual disability. Interestingly, in all ten individuals speech delays were reported. Two individuals were non-verbal, and three other individuals had a developmental language disorder diagnosis (mixed expressive/receptive language disorder in two individuals, and expressive language disorder in one). One of these latter three individuals was also reported with verbal dyspraxia. In addition, five individuals were reported to have nasal speech, and one individual was reported with persistent stuttering.

**Figuer 1:**
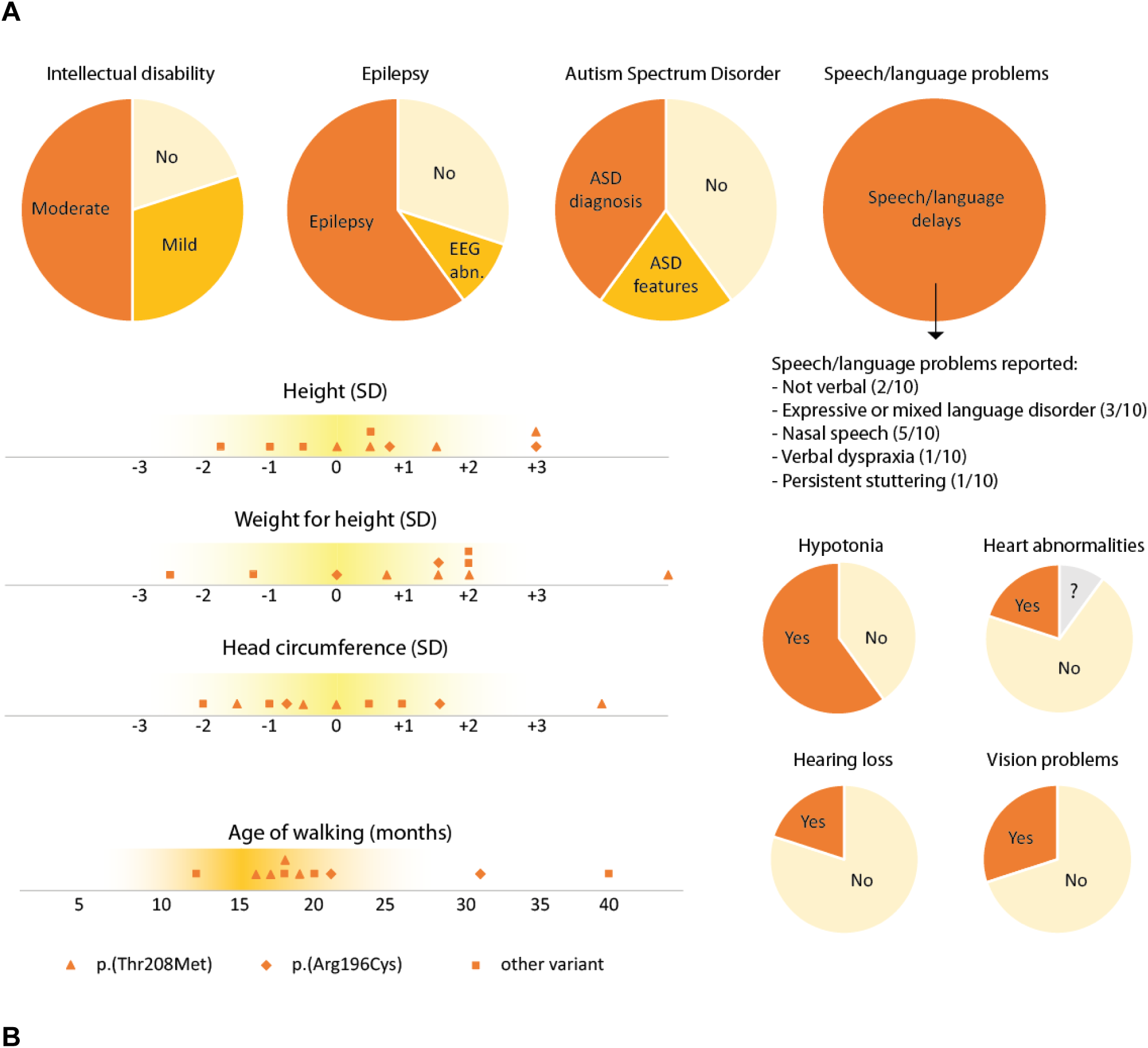
Clinical features reported in individuals with *WDR5* variants. *Photographs not available in this preprint*. A: Graphical overview of clinical features reported in ten individuals with WDR5 missense variants. Growth parameters are shown as standard deviations to the mean for a certain age. All graphs include data for all ten individuals (N=10). ‘EEG abn.’ = EEG abnormalities. B: Facial images of seven individuals with a heterozygous WDR5 variant (photographs not available in this preprint). Several overlapping facial features are seen, such as a bulbous nasal tip, low-set, posteriorly rotated and/or dysplastic ears, ptosis and thin upper lip vermilion. In addition to this, two individuals have severe micrognathia, a small mouth and down-slanting palpebral fissures.

All but one of the individuals with *WDR5* variants had motor development delays, with the age of first steps ranging from 12-40 months. Hypotonia was reported in six individuals. Concerning the behavioural phenotype, four individuals had an autism spectrum disorder (ASD) diagnosis, and two individuals were diagnosed with Attention Deficit Hyperactivity Disorder (ADHD).

Six out of the ten individuals were diagnosed with epilepsy, with different forms of presentations varying from absence seizures to refractory generalized myoclonic epilepsy. In four of these six individuals, the disorder was only present in childhood and medication could be discontinued at a later age. A brain MRI scan was performed for seven individuals, showing different types of abnormalities in three of these scans: mild ventricular dilatation with thinning of the posterior corpus callosum in one individual, subtle grey matter heterotopias in another individual, and periventricular gliosis in the third individual with an abnormal MRI result.

Individuals with *WDR5* variants showed divergent growth parameters. One individual had macrocephaly (head circumference ≥+2 SD) and another had microcephaly (≤-2 SD). Two individuals had tall stature (≥+2 SD). A weight of +2 SD or more (for height) was seen in four individuals in total, including all three adult individuals in our study. One individual had a low weight (≤-2 SD). No clear correlation between height, weight and head circumference was observed (table S1), with the exception of one individual which showed a striking pattern of progressive overgrowth (height +3 SD, weight +5 SD, head circumference +4 SD at adult age) and another individual with a milder generalized overgrowth phenotype (height +3 SD, weight +1.5 SD, head circumference +1.5 SD). Different abnormalities of the skeleton and limbs were present in a subset of individuals. Bilateral clubfeet were reported in one individual and another individual had hemihypertrophy of the left leg. One individual was reported with a hemivertebra of L5 and kyphosis (possibly secondary to the hemivertebra), and another individual had scoliosis. Two individuals had single palmar creases. In two individuals, heart abnormalities were reported: cardiac arrhythmias and decompensated heart failure requiring surgery in one individual, and left ventricular non-compaction cardiomyopathy in another individual. Three individuals were reported with frequent infections of the ears and/or airways. One individual had strabismus, another individual had amblyopia and hyperopia with astigmatism.

Significant facial dysmorphisms were noted in only a subset of individuals. When comparing facial features of seven individuals in our cohort, overlapping facial features included a bulbous nasal tip, low-set, posteriorly rotated and/or dysplastic ears, ptosis and thin lip vermilion (Figure 1B). Two individuals in our cohort had distinct facial features compared to the others, with severe micrognathia (requiring tracheostomy in one), a small mouth and prominent down-slanting palpebral fissures. These two individuals both had conductive hearing loss too, a feature not reported in any of the other individuals. Clinical features reported in individuals in our cohort are described in more detail in table S1.

### *In silico* analysis of variants

The missense variants in our cohort are all located within or flanking the fourth and the fifth WD40 domain of WDR5, with each WD40 domain encoding one ‘propeller’ of the three-dimensional WDR5 structure (Figure 2A and 2B). All missense variants in our study were absent from the gnomAD database^19^. We used *in silico* prediction programs to evaluate likely pathogenicity for the six different missense variants. The resulting scores are summarised in Table 1. All CADD scores were above 22, while SIFT and Polyphen-2 predicted three out of the six variants to be pathogenic: p.(Ala169Pro), p.(Arg196Cys) and p.(Thr208Met).

**Figuer 2.**
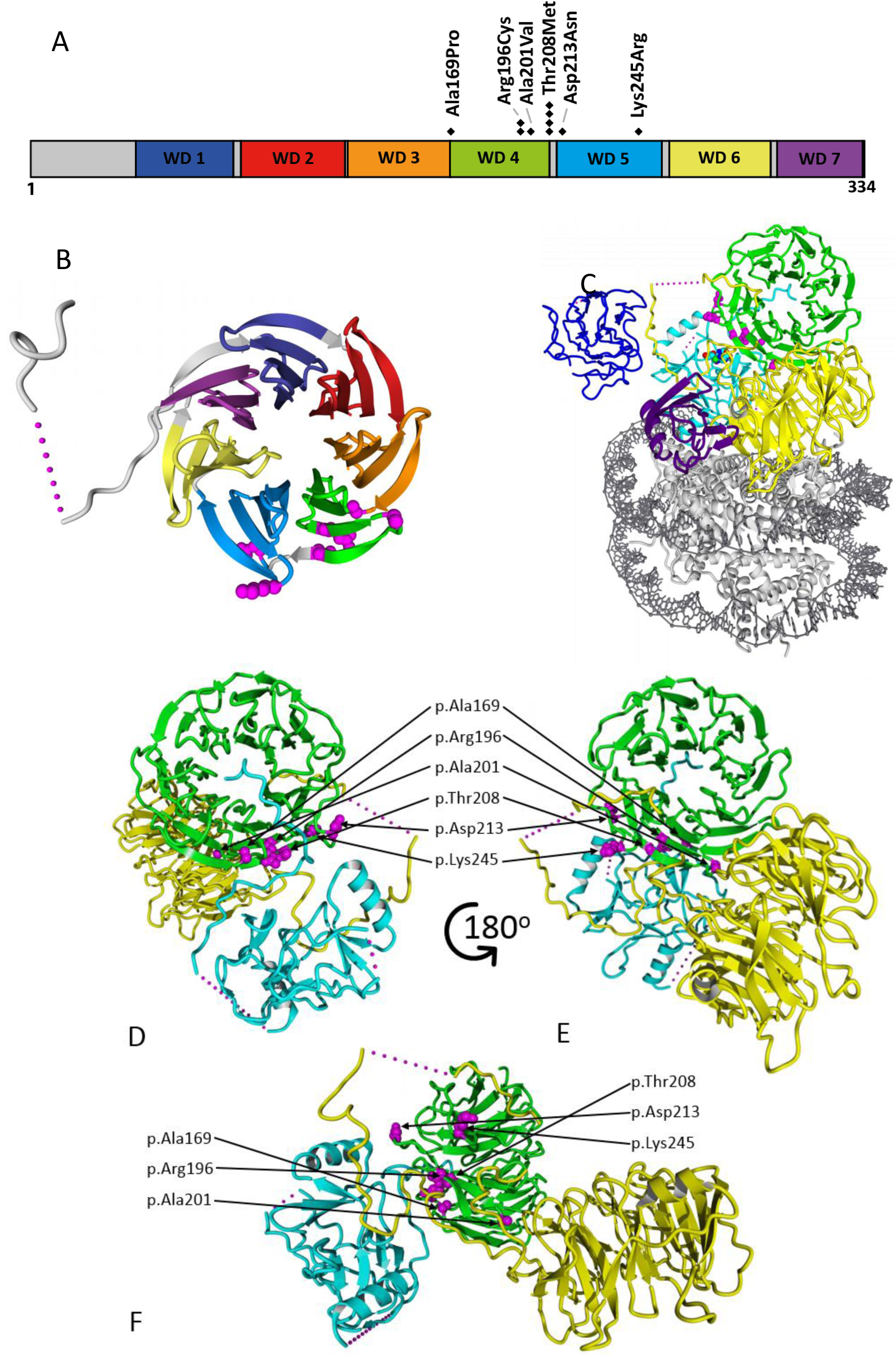
A: Linear structure of WDR5 protein (334 amino acids) with the seven different WD40-domains and all identified missense variants shown; in total six different missense variants were found in ten individuals, as one variant (p.(Thr208Met)) was found in four unrelated individuals and another variant (p.(Arg196Cys)) was found in two unrelated individuals. B:Three-dimensional visualization of WDR5 (PDB 2GNQ), locations of the amino acids involved in missense variants are shown with magenta balls. Colours of the different domains match with the colours used in panel A. C: WDR5 (green) is shown as part of the core MLL1 complex, with RbBP5 (yellow), ASH2L (blue), DPY30 (purple) and KMT2A (cyan) (PDB:6KIV). The nucleosome is shown in grey. The locations of the amino acids affected in patients identified in this study are shown with balls (magenta). D, E, F: WDR5 (green, p.33-332) is shown together with RbBP5 (yellow, p.1-380) and KMT2A (cyan, p.3764-3969), as part of the core MLL1 complex (PDB:6KIV). The locations of the amino acids affected in patients identified in this study are shown with balls (magenta), from three different angles: facing the WIN site (D), the WBM site (E) and a side between WIM and WBM sites (F).

### Three-dimensional structure analysis

Using three-dimensional structure analysis, we determined that the residues affected by all the identified missense variants cluster together on one side of the WDR5 surface, suggesting that these variants may disrupt specific interactions with other proteins (Figure 2B). Different surfaces of WDR5 are known to mediate interactions with different proteins, thereby linking them together. In previous co-precipitation experiments with WDR5 and short fragments of proteins from complexes in which WDR5 is involved, two distinct binding sites were identified: the ‘WDR5-interacting’ (WIN) site^39-41^ and the ‘WDR5-binding motif’ (WBM) site^41,42^, located on opposite sides (often referred to as ‘top’ and ‘bottom’, respectively) of the protein. The variants found in our study are not located in the vicinity of these well-studied binding locations.

However, recently published cryo-electron microscopy 3D structures of the core MLL1 and MLL3 complexes revealed a region located between the WIN and WBM binding sites, that is involved in the interaction with RbBP5 and/or histone-lysine methylase (KMT) enzymes in these complexes^36^. Five out of the six variants that we identified map within this RbBP5/KMT interaction region. Based on the 3D structure analysis of the MLL1 and MLL3 complexes, p.(Ala169Pro) and p.(Asp213Asn) are predicted to affect the WDR5 interaction with KMT enzymes, and p.(Ala201Val) and p.(Thr208Met) are predicted to affect the interaction with the RbBP5 enzymes, while p.(Arg196Cys) most likely influences the interaction with both enzymes (Figure 2C-F). The effects of the p.(Lys245Arg) variant cannot be predicted using the currently available 3D structures. A detailed description of the predicted effects of all variants, from the perspective of the structural modelling analyses, is provided in Supplementary Note 1.

## Discussion

We identified rare *de novo* missense variants in *WDR5*, a gene that encodes a core chromatin regulator and is extremely intolerant for variation in the population. We clarify the molecular and phenotypic consequences of these rare variants which define a novel Mendelian disorder. By using the GeneMatcher database and international collaborations, we collected clinical data on a cohort of ten individuals with a *de novo* variant in *WDR5*. We studied all variants in three-dimensional conformations of WDR5 in interaction with COMPASS complex family subunits, and showed that the missense variants are predicted to affect important binding sites of WDR5. By combining data from these clinical and *in silico* approaches, we can confidently link *WDR5* to a neurodevelopmental disorder with a broad spectrum of associated features, further confirming that proteins in the COMPASS complex family are important contributors to neurodevelopment.

All individuals in our cohort had a heterozygous *de novo* missense variant in *WDR5*: six different missense variants were found, affecting amino acids on one surface of the WDR5 protein. The recurrence of the p.(Thr208Met) and p.(Arg196Cys) variant in four and two unrelated individuals, respectively, points to the presence of ‘hotspots’ for recurrent variants in *WDR5*. Three different missense variants in our cohort, p.(Ala169Pro), p.(Arg196Cys) and p.(Thr208Met), were found to locate at adjacent amino positions in the three-dimensional structure of WDR5. The fact that all identified variants are missense variants, the recurrence of specific variants in unrelated individuals and the spatial clustering of the variants on the protein surface of WDR5, all suggest that specific pathogenic mechanisms that might be at play are not just loss-of-function or haploinsufficiency.

To the best of our knowledge, truncating variants (e.g. frameshift or nonsense variants) in *WDR5* have not been identified so far: not in our cohort or any disease cohort in literature, nor in healthy individuals (e.g. in the gnomAD or TOPMED database). According to sequencing data from the gnomAD database, *WDR5* is extremely intolerant for both missense and loss-of-function variation. The gene has a LOEUF score of 0.124, which is well within the first decile of most highly constrained genes against loss-of-function^19^. In contrast to the absence of truncating variants, heterozygous chromosomal microdeletions encompassing the whole *WDR5* gene have been reported; the Decipher database lists eleven heterozygous deletions that include WDR5^43^. This means that haploinsufficiency for *WDR5* is compatible with life, but it is unclear how the loss of WDR5 contributes to specific phenotypes found in individuals with these deletions, as all deletions are larger than 3 Mb and encompass many other genes as well.

Analysis of the missense variants in a three-dimensional structure of WDR5 in the context of the MLL1 or MLL3 complex, revealed that all but one were located at amino-acid positions that are important for binding of WDR5 with other proteins of these complexes. Interestingly, analysis of the intolerance landscape of the *WDR5* gene in the three-dimensional structure of the encoded protein shows that WDR5 is generally intolerant to missense variants, but that residues interacting with other proteins are most intolerant for normal variation (Figure S1). For five out of six missense variants, we predict that the corresponding amino-acid change disrupts the interaction of WDR5 with the MLL1/MLL3 complex subunits RbBP5 and with KMT2A/C simultaneously or separately. WDR5 is a crucial core protein within the COMPASS complex family; it is essential for complex assembly and activity^44,45^. Based on 3D analysis of the MLL1 and MLL3 complexes, it seems that differently composed COMPASS complexes make use of different interaction surfaces of WDR5. Some variants might therefore disrupt interactions in only one specific complex. As the core function of the WDR5 protein seems to be to act as an ‘adapter’ protein and form links between different molecules, disruption of protein-protein interactions within the complex might have important effects on complex activity.

However, it is important to take into account that the extensive and detailed three-dimensional structures that we used for these analyses are only available for the MLL1/3 complex, and not for all other complexes and interactions in which WDR5 is involved. Therefore, it remains unclear whether the predicted disruptive effects on WDR5 interactions are specific to those with RbBP5/KMT and MLL1/3, or whether interactions with other molecules might also be disturbed. For the p.(Lys245Arg) variant, we were not able to predict a possible pathogenic mechanism analysing available three-dimensional structures. One hypothesis could be that the variant affects a so far uncharacterized interaction site with RbBP5 or KMT2A/C, as a comparison of available three-dimensional structures between human MLL1 and yeast COMPASS complex suggest even more extensive interaction surfaces between WDR5 and histone methylases (Figure S2). Another hypothesis is that the p.(Lys245Arg) variant affects the interaction with other molecules that are not involved in the MLL1/MLL3 complex. Lastly, although the *de novo* occurrence of the p.(Lys245Arg) and the phenotypic similarity to the rest of the cohort suggest pathogenicity, the possibility that this is a benign variant without any effect on WDR5 function cannot be excluded.

The individuals in our study showed a broad range of clinical features: neurodevelopmental phenotypes with several additional abnormalities. All individuals had developmental delays, with mild or moderate intellectual disability being present in the majority. Speech and language problems were a prominent feature, as was epilepsy. Abnormal height, weight and head circumferences were frequently seen, and two individuals had a remarkable distinct facial phenotype with severe micrognathia, a small mouth and down-slanted palpebral fissures. When comparing the phenotypes corresponding to different variants in our study, no clear genotype-phenotype correlation was established. Even in four individuals with the exact same p.(Thr208Met) variant a different clinical presentation was seen: e.g. borderline vs. moderate intellectual disability, normal growth parameters vs. a generalized overgrowth phenotype, and variability in the presence of additional phenotypic features. Altogether, *WDR5*-associated disorder can be characterized as a neurodevelopmental disorder with variable expressivity of associated features. This is in line with other disorders caused by variants in genes encoding COMPASS complex family subunits, such as Kabuki syndrome (caused by variants in *KMT2D* or *KDM6A*)^46^, Kleefstra syndrome type 2 (caused by variants in *KMT2C*)^47^ and the neurodevelopmental disorder associated with *SETD1A* loss-of-function variants^48-50^. In all these COMPASS complex-associated disorders a variable spectrum of associated features can be present in varying degrees of severity, including intellectual disability, speech and language delays, autism spectrum disorders, epilepsy and abnormal growth parameters^46-50^.

This study represents the first characterization of a Mendelian disorder associated with germline variants in WDR5, and was initiated after the report of a *de novo* WDR5 variant in a child with a speech disorder^21^. It is worth mentioning that one additional *de novo* variant in *WDR5* is reported in the literature: a p.(Lys7Gln) variant, found in a child with a conotruncal heart defect with a right aortic arch^51^. This missense variant is located in the N-terminal tail of WDR5, an intrinsically disordered region of the protein (not available for three-dimensional protein structure analysis), which is not involved in the beta-propeller structure of WDR5, and has been shown to be dispensable for COMPASS complex assembly^52^. A study in *Xenopus tropicalis* shows that this p.(Lys7Gln) variant might interfere with the ability of WDR5 to localize to the bases of left-right organizer cilia, independent from the H3K4-methylation-related functions of WDR5^53^. As p.(Lys7Gln) is located in a different region of the WDR5 protein compared to the variants found in our study, and since complete phenotypic details are not available for this individual, it is currently unclear whether this reported individual has the *WDR5-*associated neurodevelopmental disorder presented in this study, or whether this specific variant gives rise to a different disorder with different pathogenic mechanisms.

All in all, with this study WDR5 can be added to the list of genes robustly associated with autosomal dominant neurodevelopmental disorders. All variants found so far are missense variants, and although they are not always predicted to be pathogenic by the commonly used prediction programs, our three-dimensional protein structure analysis showed that it is very likely that five out of the six found missense variants disturb important protein-protein interactions. Future studies are needed to confirm that these *in silico* observations in 3D structures indeed affect protein-protein interactions between WDR5 and RbBP5 and KMT enzymes, and what the downstream consequences are on H3K4 methyltransferase activity. But taken together, with our study we implicate *WDR5* variants in an autosomal dominant disorder associated with intellectual disability, speech and language delays, epilepsy and autism spectrum disorder, thereby highlighting the role of COMPASS complex family protein disruptions in neurodevelopmental disorders.

## Supporting information

COI disclosure form

## Data Availability

Figure 1B and Table S1 are available from the corresponding author on request.

## Data availability

Figure 1B and Table S1 are available from the corresponding author on request.

## Declaration of interests

F.M. is a full time employee at GeneDx, Inc. The authors declare no other conflicts of interest.

## Acknowledgements

We thank all included individuals and their families for their contribution to this research project.

This work was generated within ITHACA: European Reference Network on Rare Congenital Malformations and Rare Intellectual Disability. Funding was provided by the Netherlands Organization for Scientific Research (NWO) Gravitation Grant 24.001.006 to the Language in Interaction Consortium (L.S.B., S.E.F., and H.G.B.), the Max Planck Society (S.E.F.) and the Netherlands Organization for Health Research and Development (ZonMw grant 91718310 to T.K.). The research of A.C., M.I. and E.R. was supported by PROGETTEO GENE (GENE = Genomic analysis Evaluation Network) founded by PROGETTI DI INNOVAZIONE IN AMBITO SANITARIO E SOCIO SANITARIO (BANDO EX DECRETO N. 2713 DEL 28/02/2018). A.R. and N.K. acknowledge funding from the European Regional Development Fund (ERDF). K.Õ. And K.R. were supported by the Estonian Research Council grants PUT355 and PRG471. The Broad Center for Mendelian Genomics (UM1 HG008900) is funded by the National Human Genome Research Institute with supplemental funding provided by the National Heart, Lung, and Blood Institute under the Trans-Omics for Precision Medicine (TOPMed) program and the National Eye Institute.

Individuals 3 and 4 in this study were part of the DDD study cohort. The DDD study presents independent research commissioned by the Health Innovation Challenge Fund [grant number HICF-1009-003]. This study makes use of DECIPHER (http://decipher.sanger.ac.uk), which is funded by Wellcome. See Nature PMID: 25533962 or www.ddduk.org/access.html for full acknowledgement.

## Notes

### Author Declarations

This study was approved by the institutional review board 'Commissie Mensgebonden Onderzoek Regio Arnhem-Nijmegen' under number 2011/188. This number refers to performing diagnostic WES. Discovery of novel syndromes/description if cases from this series can be taken as such. All necessary patient/participant consent has been obtained and the appropriate institutional forms have been archived.

