## Supplemental Data for "A clustering of missense variants in the crucial chromatin modifier WDR5 defines a new neurodevelopmental disorder"

### Supplementary Data

##### Figure S1 MetaDome intolerance visualization of WDR5

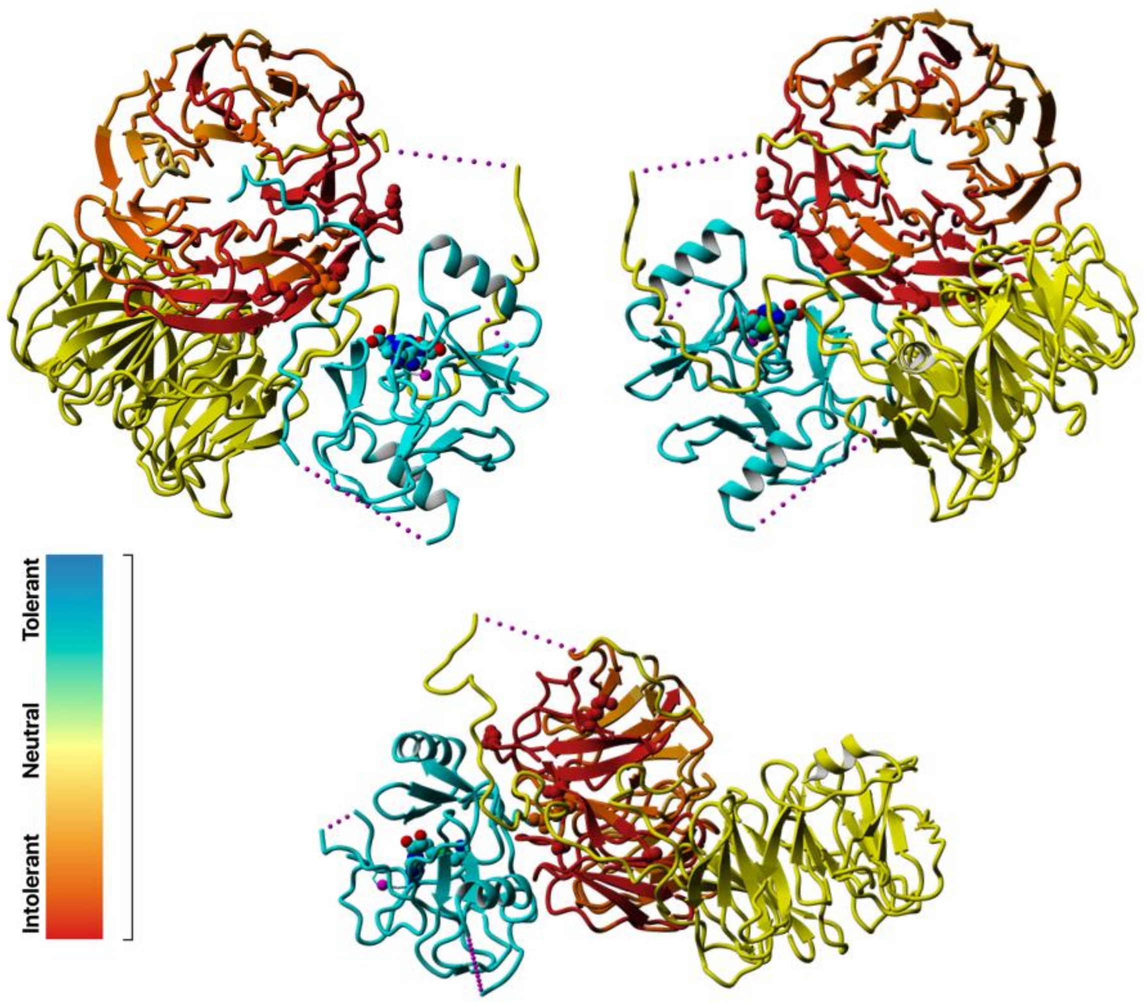

WDR5 is coloured in line with the MetaDome tolerance scale shown. RbBP5 is shown in yellow and KMT2A in cyan (PDB:6KIV). As can be seen in this figure, WDR5 is generally intolerant to missense variants, but WDR5 amino acids that are known to interact with other proteins are most intolerant (darker red).

**Figure S2: Comparison of the core human MLL1 with the yeast COMPASS complexes**

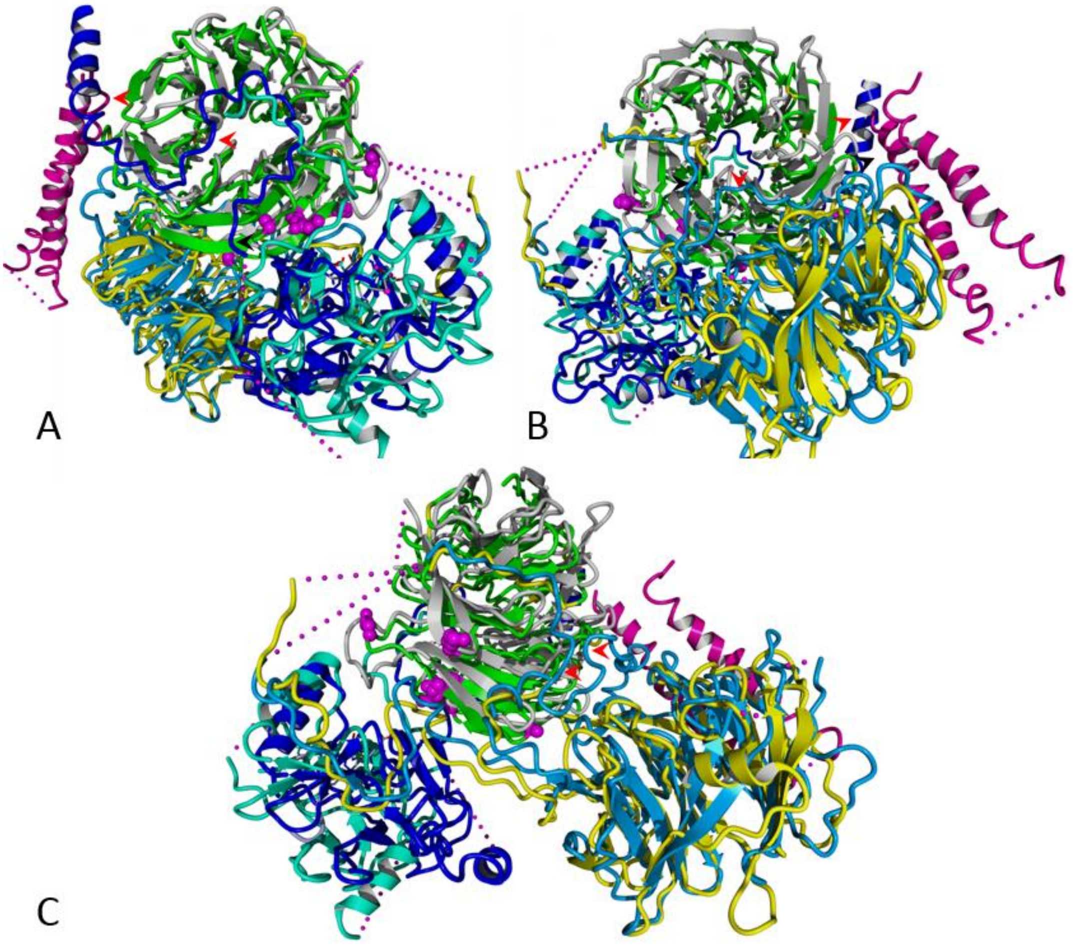

The alignment of human WDR5 in complex with RbBP5 and KMT2A/MLL1 from the core MLL1 complex (PDB:6KIV) with homologues of the yeast COMPASS complex (PDB:6UH5) is shown: WDR5 (green, p.33-332) with its homologue Swd3 (grey, p.16-326); RbBP5 (yellow, p.1-380) with its homologue Swd1 (light blue, p.1-435); KMT2A/MLL1 (cyan, p.3764-3969) with yeast homologue Set1c (dark blue, p.819-999). Additionally, yeast Spp1 (purple) is shown. The Spp1 homologue is not present in human COMPASS family complexes. The locations of the amino acids that are affect in patients identified in this study are shown with balls (magenta). Three different angles are shown: WDR5 faced from the WIN site (A), from the WBM site (B), and from the side between WIN and WBM (C).

The human core COMPASS/COMPASS family complexes (eg., MLL1) are highly conserved and have a structure similar to the yeast COMPASS complex. Because the yeast COMPASS complex proteins in the 3D model are more complete, substantially more extensive interaction of the RbBP5 and KMT2A/MLL1 homologues with WDR5 homologues can be observed (red arrows). Additionally, another interaction site of the WDR5 homologue is observed with a Spp1 protein

These 3D modelling data, in addition to the high conservation level and low tolerance to the missense and LoF variants in the general population, suggest that also human WDR5 may have significantly more extensive interaction surfaces within COMPASS family complexes and other chromatin-remodelling complexes.

#### Supplementary note 1: Detailed description and visualization of the predicted effect of identified WDR5 variants

##### *p.(Ala169Pro)*

| Wild type residue role | Effect of the residue substitution |
|---|---|
| Ala169 is located in a turn from the third to fourth WDR5 beta-propeller. Despite the fact that the Ala169 is located in close proximity to the KMT2A/MLL1, and KMT2C/MLL3 interaction sites, it does not directly interact with the KMT enzymes. | Change from the alanine to a larger proline at this position is predicted to result in a local backbone change, because of the rigid sidechain of the proline. This change is predicted to disturb the flexibility and local structure of WDR5, which will disrupt the binding to the KMT enzymes. |

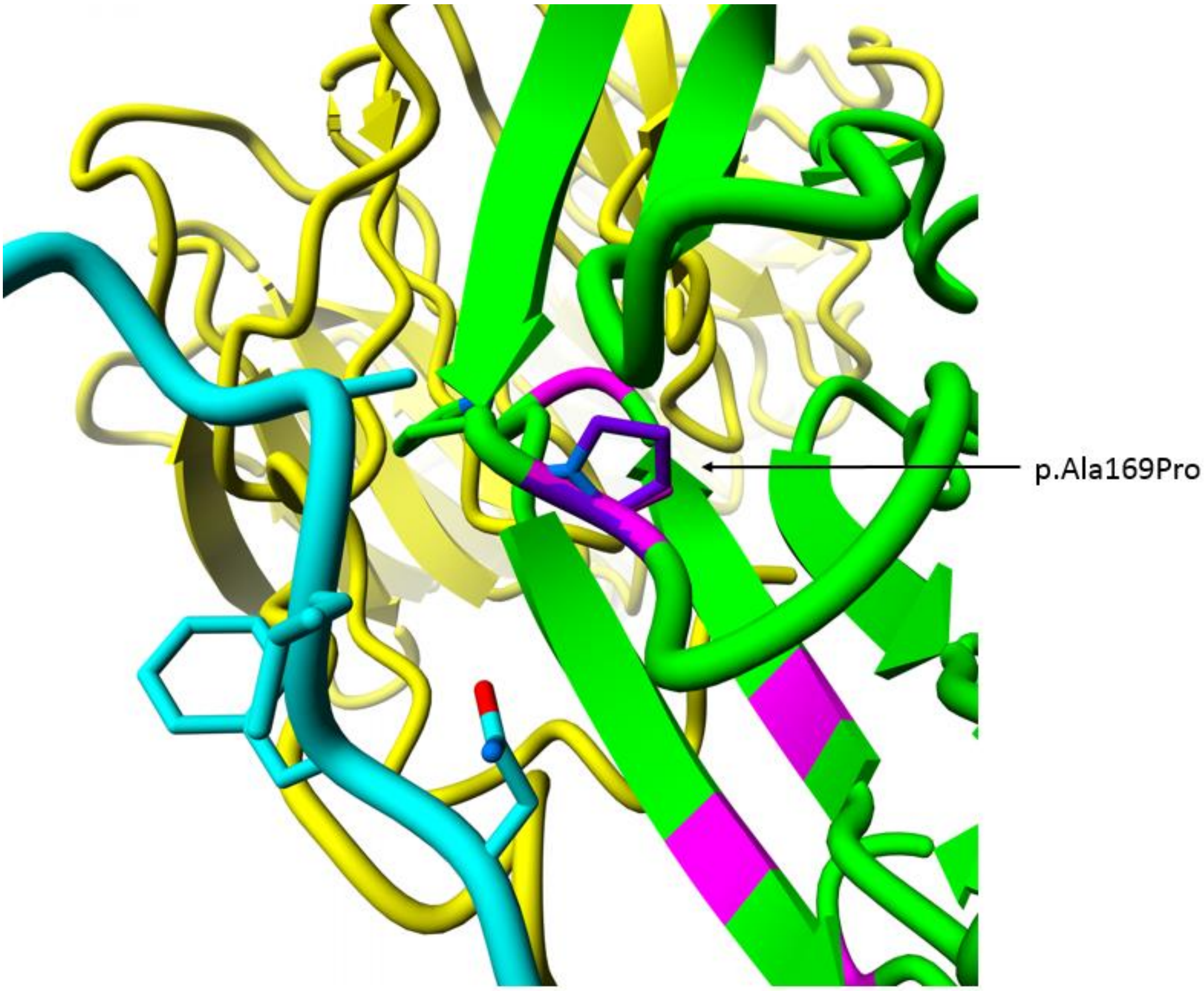


***Figure SN1:*** *WDR5 (green) interaction with KMT2A/MLL1 (cyan) and RbBP5 (yellow), are shown from the core MLL1 complex (PDB:6KIV). The mutated aminoacid and nearby aminoacids are shown with sticks. The wild type alanine at the position p.169 is colored in magenta and the mutated proline in purple.*

***p.(Arg196Cys)***

| **Wild type residue role** | **Effect of the residue substitution** |
|---|---|
| Arg196 is located on the WDR5 lateral surface for interaction with RbBP5 and KMT2A enzymes. Arg196 interacts with Asn3779 in the KMT2A protein and Phe332 in the C-term tail of RbBBP5 but has no visible interactions with the KMT2C protein. | Cysteine is a much smaller residue and does not have a charge. Therefore, a change from the arginine to cysteine at this position would result in a loss of the hydrogen-bond with Asn3779 in KMT2A, as well resulting in an empty pocket between the WDR5, KMT2A and RbBBP5 interaction surfaces, which would lead to a loss of packing interactions and disruption of the interactions between the proteins. |

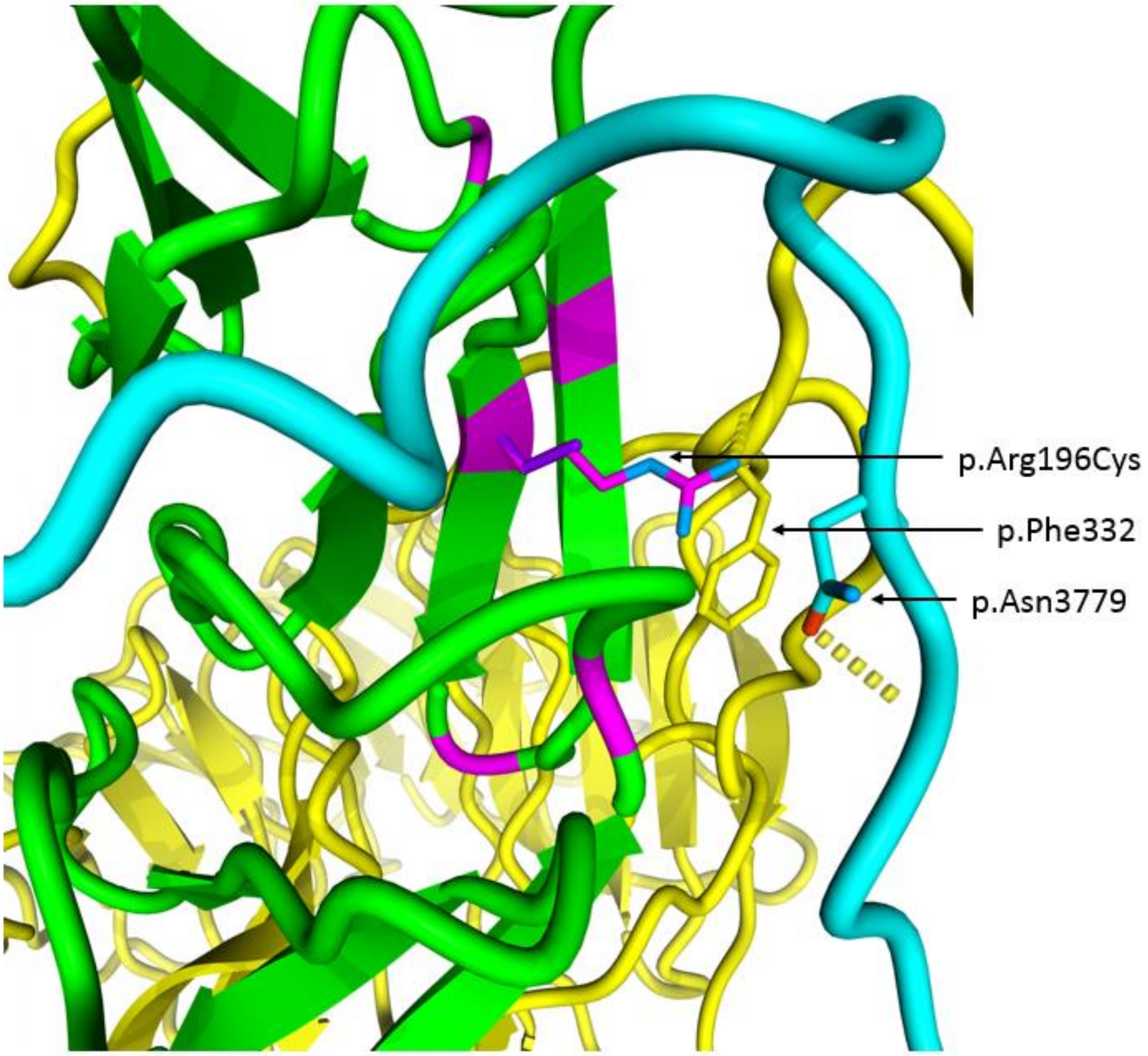


***Figure SN2:*** *WDR5 (green) interaction with KMT2A/MLL1 (cyan) and RbBP5 (yellow), are shown from the core MLL1 complex (PDB:6KIV). The mutated aminoacid and nearby aminoacids are shown with sticks. The wild type arginine at the position p.196 is colored in magenta and the mutated cysteine in purple.*

***p.(Ala201Val)***

| **Wild type residue role** | **Effect of the residue substitution** |
|---|---|
| Ala201 is located on the WBM surface of WDR5 and interacts with RbBP5. It is located in clear proximity to Arg56 and Leu54 in the RbBP5 protein. | Despite the fact that valine is also small and non-polar, it has a bigger sidechain than alanine. Therefore, change to a valine at this position could affect the interaction with RbBP5 because of the change of the interaction surface. |

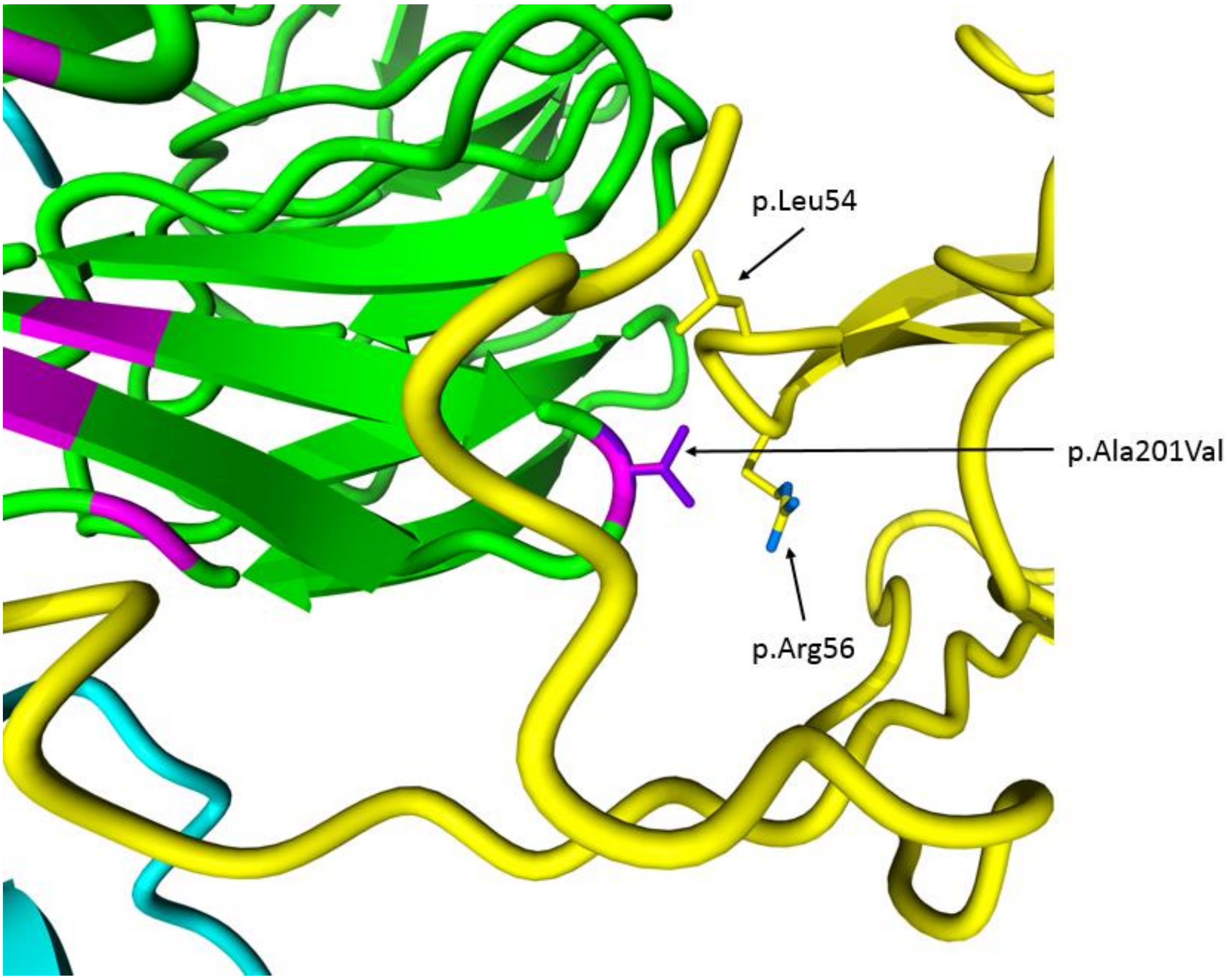


***Figure S3:*** *WDR5 (green) interaction with KMT2A/MLL1 (cyan) and RbBP5 (yellow), are shown from the core MLL1 complex (PDB:6KIV). The mutated aminoacid and nearby aminoacids are shown with sticks. The wild type alanine at the position p.201 is colored in magenta and the mutated valine in purple.*

***p.(Thr208Met)***

| Wild type residue role | Effect of the residue substitution |
|---|---|
| Thr208 in WDR5 interacts with several RbBP5 C-term tail amino acids (Ala331, Pro334). Additionally, it makes a hydrogen-bond with a backbone of Ala331 in RbBBP5. | Methionine has a substantially bigger size than threonine and is not able to form the hydrogen-bond with RbBP5 Ala331. Therefore, a change to methionine at this position is expected to disrupt the WDR5 interaction interface with RbBP5. |

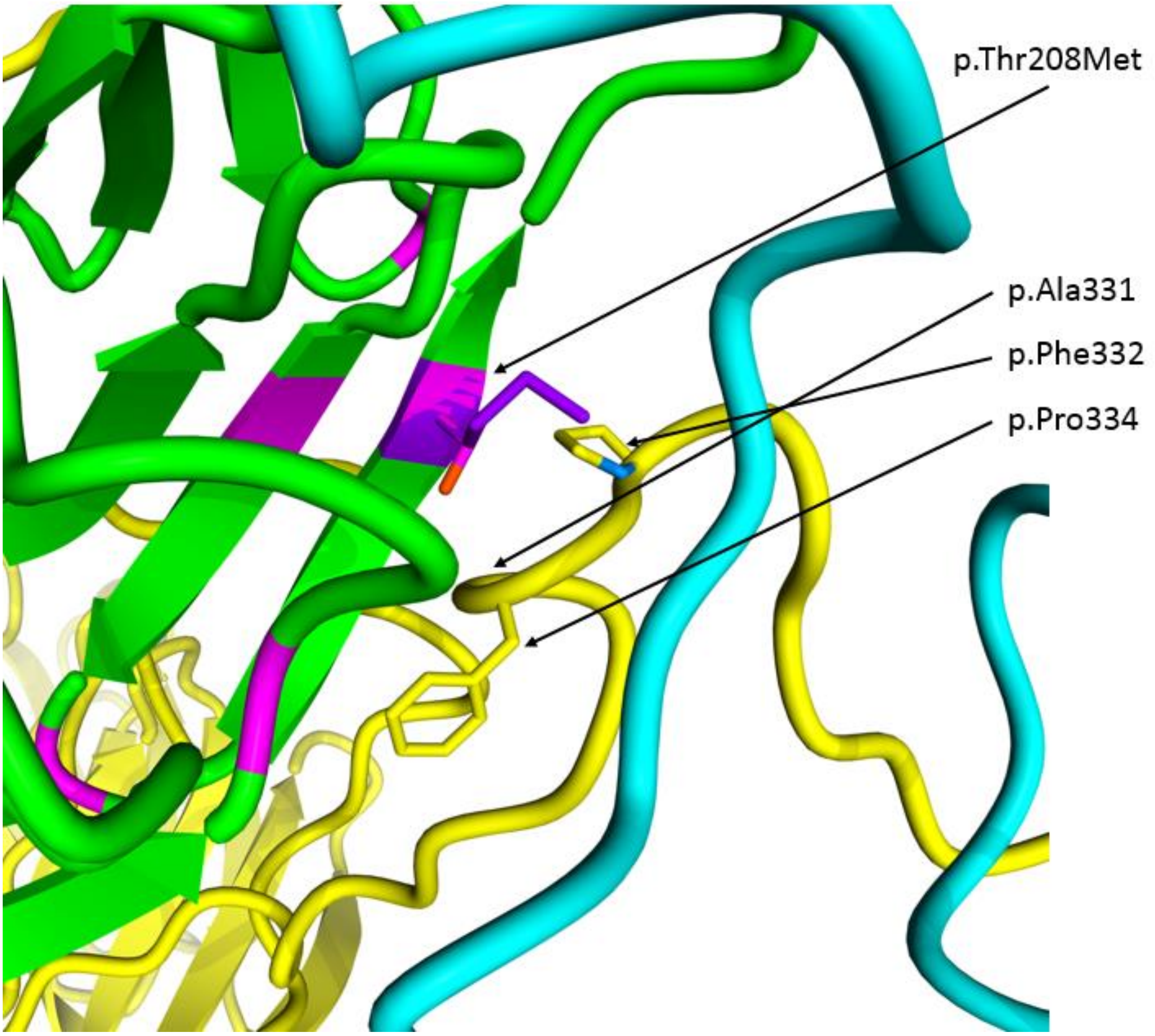


***Figure S4:*** *WDR5 (green) interaction with KMT2A/MLL1 (cyan) and RbBP5 (yellow), are shown from the core MLL1 complex (PDB:6KIV). The mutated aminoacid and nearby aminoacids are shown with sticks. The wild type threonine at the position p.208 is colored in magenta and the mutated methionine in purple.*

***p.(Asp213Asn)***

| Wild type residue role | Effect of the residue substitution |
|---|---|
| Asp213 is located in a WDR5 hydrophylic loop, which is involved in the interaction with the KMT enzymes. Although located distantly, it may interact with a positively charged KMT2A Lys3878, because the lysine has a highly flexible sidechain.<br><br>Additionally, Asp213 forms a hydrogen-bond with Asn235 in WDR5. | Change to the aspartate would result in a similar amino acid with similar size, although the negative charge of the aspartic acid would be lost. The hydrogen bond with WDR5 Asn235 would be lost due to this change, which may disrupt the stability and position of the loop, and, therefore, affect interaction with the KMT enzymes. Additionally, it can lose interactions with positively charged KMT residues. However, the exact effect of the variant is unknown. |

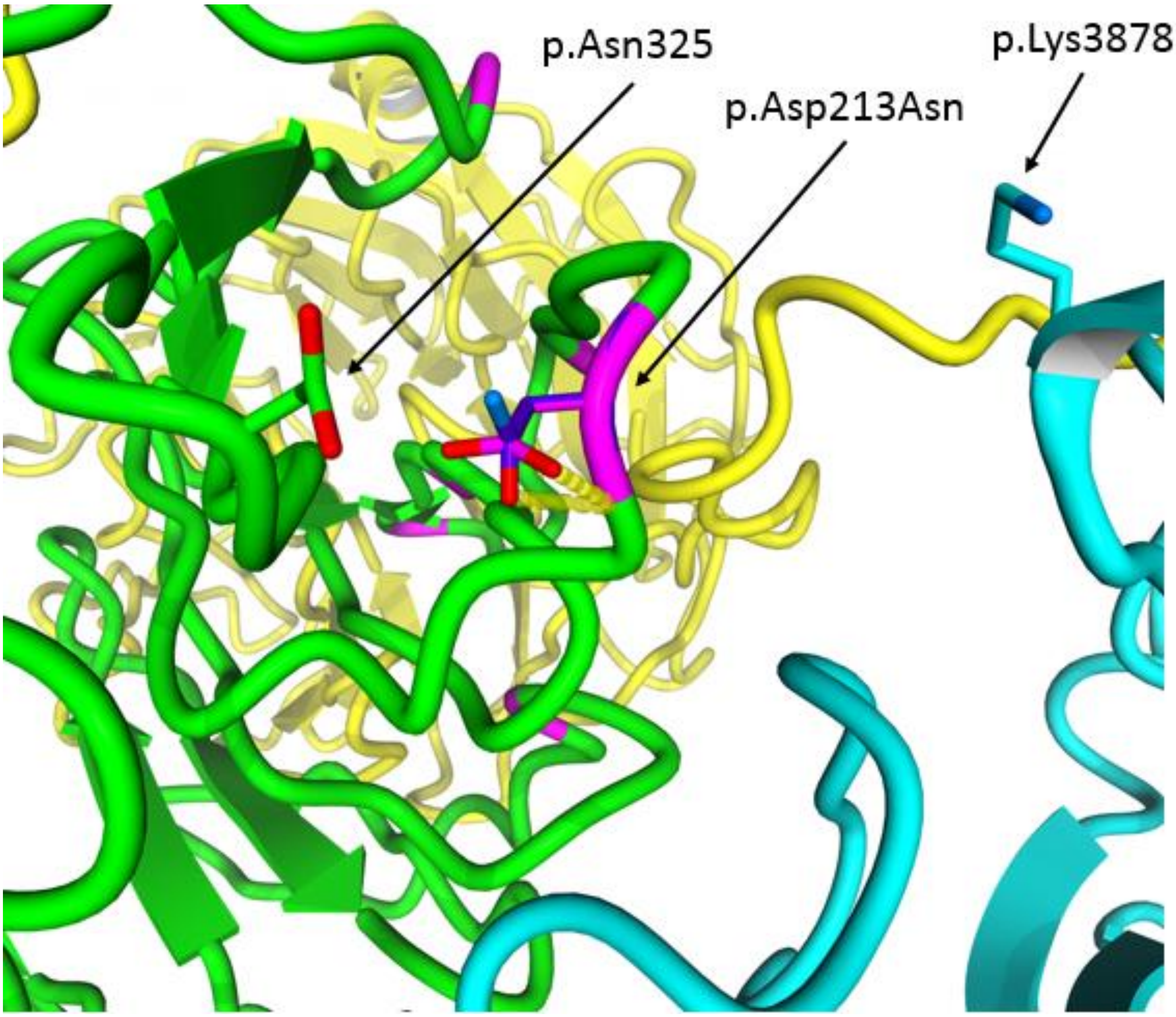


***Figure S5A:*** *WDR5 (green) interaction with KMT2A/MLL1 (cyan) and RbBP5 (yellow) are shown from the core MLL1 and MLL3 complexes (PDB:6KIV and 6KIW, respectively). The mutated aminoacid and nearby aminoacids are shown with sticks. The wild type aspartic acid at the position p.213 is colored in magenta and the mutated aspartate in purple.*

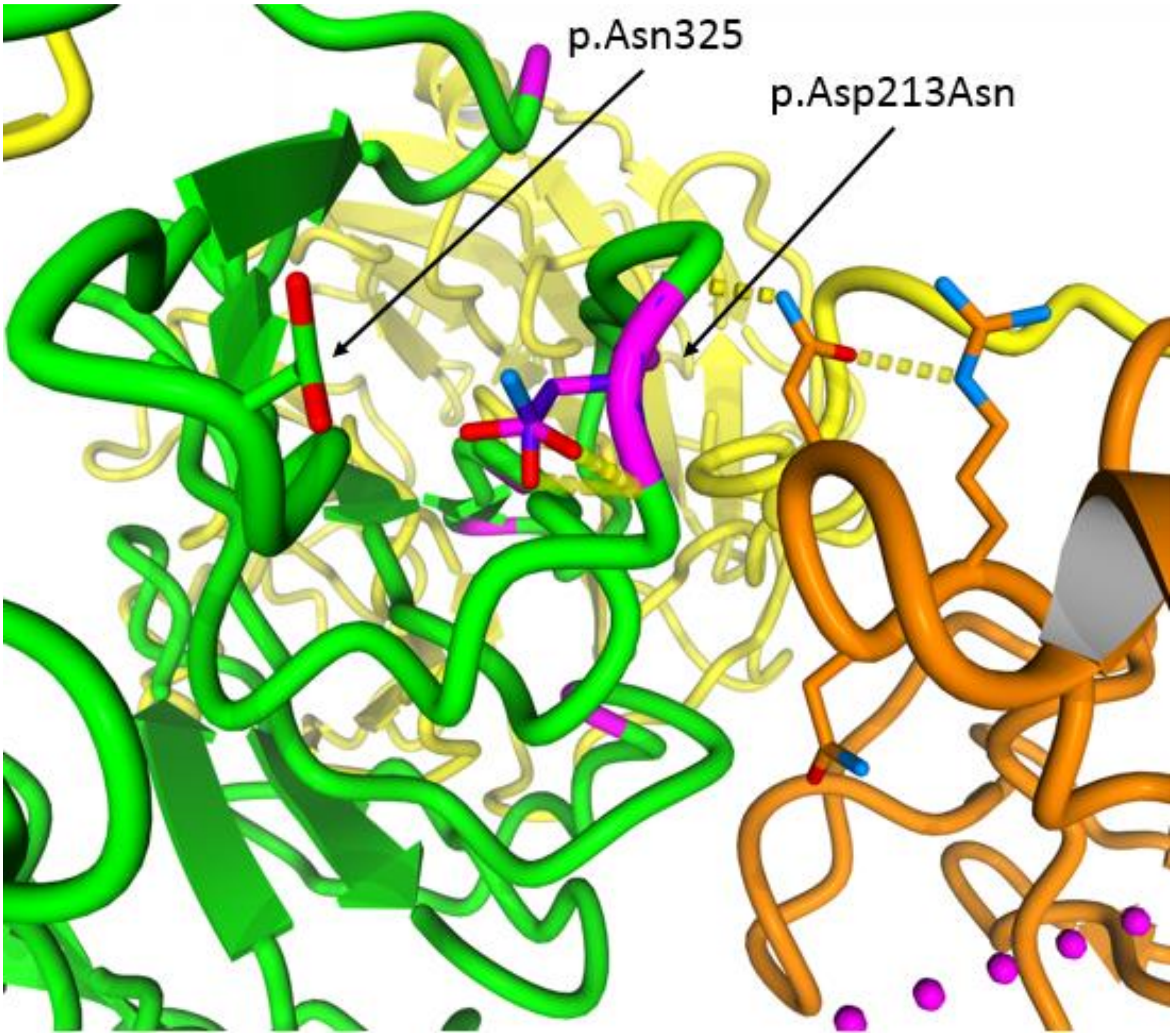


***Figure S5B:*** *WDR5 (green) interaction with KMT2C/MLL3 (orange) and RbBP5 (yellow) are shown from the core MLL1 and MLL3 complexes (PDB:6KIV and 6KIW, respectively). The mutated aminoacid and nearby aminoacids are shown with sticks. The wild type aspartic acid at the position p.213 is colored in magenta and the mutated aspartate in purple.*

***p.(Lys245Arg)***

| **Wild type residue role** | **Effect of the residue substitution** |
|---|---|
| Lys245 located in a position that is a significant distance from the site of interaction with RbBP5 and KMT enzymes and is not known to be involved in a protein interaction. | A change from lysine to arginine at this position, would result in a similar amino acid by charge and flexibility of the side-chain with minimal effect on protein structure or interactions. Even though arginine is slightly larger, the effect of this variant is not clear. |

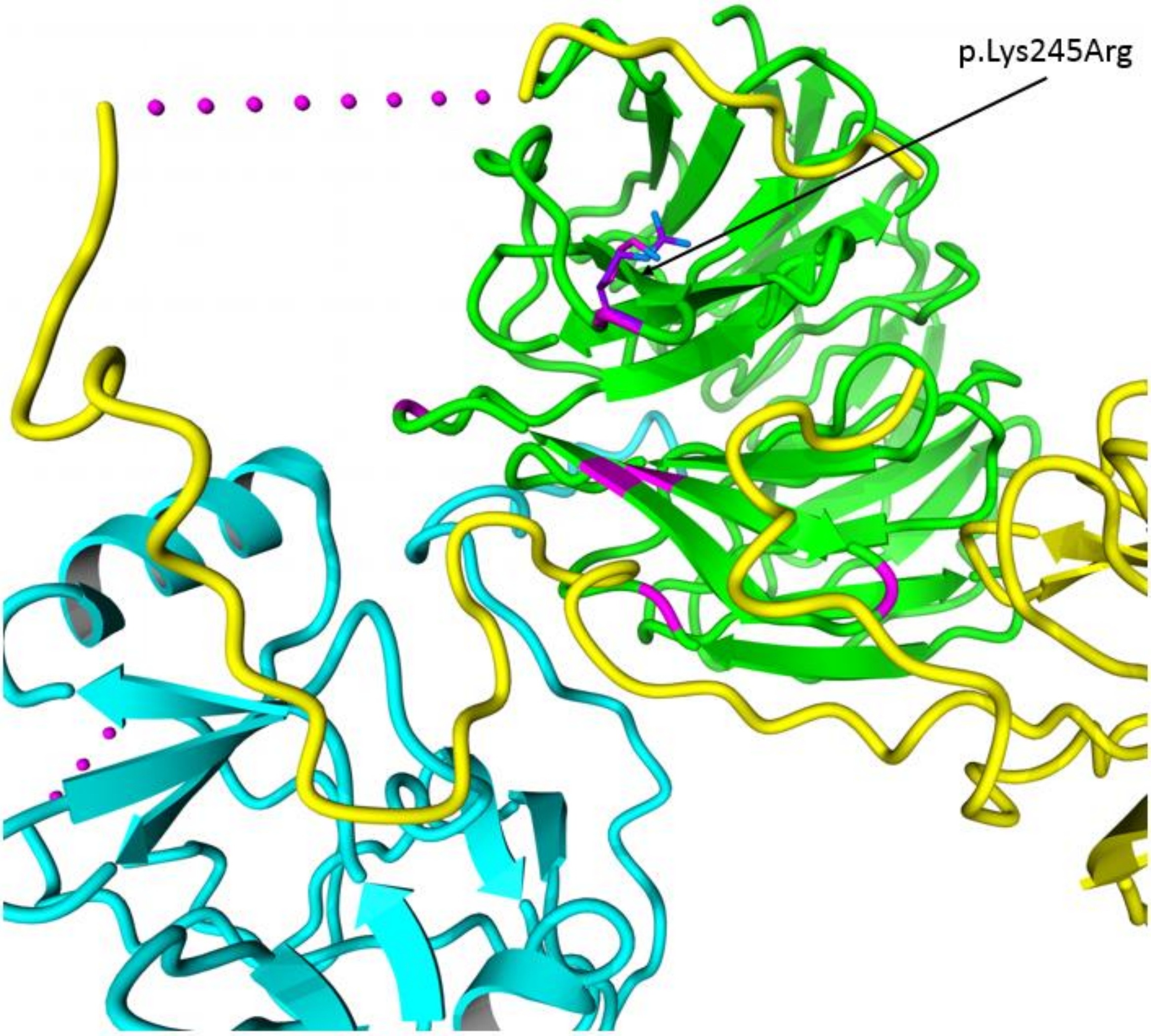


***Figure S6:*** *WDR5 (green) interaction with KMT2A/MLL1 (cyan) and RbBP5 (yellow), are shown from the core MLL1 complex (PDB:6KIV). The mutated aminoacid and nearby aminoacids are shown with sticks. The wild type lysine at the position p.245 is colored in magenta and the mutated arginine in purple.*
