## Supplementary material for "A clustering of missense variants in the crucial chromatin modifier WDR5 defines a new neurodevelopmental disorder": COI disclosure form

### ICMJE DISCLOSURE FORM

| | |
|---|---|
| **Date:** | 10/26/2021 |
| **Your Name:** | Lot Snijders Blok |
| **Manuscript Title:** | A clustering of missense variants in the crucial chromatin modifier WDR5 defines a new neurodevelopmental disorder |
| **Manuscript Number (if known):** | MEDRXIV/2021/265518 |

In the interest of transparency, we ask you to disclose all relationships/activities/interests listed below that are related to the content of your manuscript. "Related" means any relation with for-profit or not-for-profit third parties whose interests may be affected by the content of the manuscript. Disclosure represents a commitment to transparency and does not necessarily indicate a bias. If you are in doubt about whether to list a relationship/activity/interest, it is preferable that you do so.

The author's relationships/activities/interests should be defined broadly. For example, if your manuscript pertains to the epidemiology of hypertension, you should declare all relationships with manufacturers of antihypertensive medication, even if that medication is not mentioned in the manuscript.

In item #1 below, report all support for the work reported in this manuscript without time limit. For all other items, the time frame for disclosure is the past 36 months.

| | | Name all entities with whom you have this relationship or indicate none (add rows as needed) | Specifications/Comments (e.g., if payments were made to you or to your institution) |
|---|---|---|---|
| **Time frame: Since the initial planning of the work** | | | |
| 1 | All support for the present manuscript (e.g., funding, provision of study materials, medical writing, article processing charges, etc.) No time limit for this item. | ☐ None<br><br>This work was generated within ITHACA: European Reference Network on Rare Congenital Malformations and Rare Intellectual Disability. Funding was provided by the Netherlands Organization for Scientific Research (NWO) Gravitation Grant 24.001.006 to the Language in Interaction Consortium (L.S.B., S.E.F., and H.G.B.), the Max Planck Society (S.E.F.) and the Netherlands Organization for Health Research and Development (ZonMw grant 91718310 to T.K.). The research of A.C., M.I. and E.R. was supported by PROGETTEO GENE (GENE = Genomic analysis Evaluation Network) founded by PROGETTI DI INNOVAZIONE IN AMBITO SANITARIO E SOCIO SANITARIO (BANDO EX DECRETO N. 2713 DEL 28/02/2018). A.R. and N.K. acknowledge funding from the European Regional Development Fund (ERDF). K.Õ. and K.R. were supported by the Estonian Research Council grants PUT355 and PRG471. The Broad Center for Mendelian Genomics (UM1 HG008900) is funded by the National Human Genome Research Institute with supplemental funding provided by the National Heart, Lung, and | |

| | | Name all entities with whom you have this relationship or indicate none (add rows as needed) | Specifications/Comments (e.g., if payments were made to you or to your institution) |
|---|---|---|---|
| | | Blood Institute under the Trans-Omics for Precision Medicine (TOPMed) program and the National Eye Institute.<br>Individuals 3 and 4 in this study were part of the DDD study cohort. The DDD study presents independent research commissioned by the Health Innovation Challenge Fund [grant number HICF-1009-003]. This study makes use of DECIPHER (http://decipher.sanger.ac.uk), which is funded by Wellcome. See Nature PMID: 25533962 or www.ddduk.org/access.html for full acknowledgement. | |
| | | | Click the tab key to add additional rows. |
| **Time frame: past 36 months** | | | |
| **2** | Grants or contracts from any entity (if not indicated in item #1 above). | ☐ **None** | |
| | | See statement above | |
| **3** | Royalties or licenses | ☐ **None** | |
| **4** | Consulting fees | ☒ **None** | |
| **5** | Payment or honoraria for lectures, presentations, speakers bureaus, manuscript writing or educational events | ☒ **None** | |

| | | Name all entities with whom you have this relationship or indicate none (add rows as needed) | Specifications/Comments (e.g., if payments were made to you or to your institution) |
|---|---|---|---|
| **6** | Payment for expert testimony | ☒ **None** | |
| **7** | Support for attending meetings and/or travel | ☒ **None** | |
| **8** | Patents planned, issued or pending | ☒ **None** | |
| **9** | Participation on a Data Safety Monitoring Board or Advisory Board | ☒ **None** | |
| **10** | Leadership or fiduciary role in other board, society, committee or advocacy group, paid or unpaid | ☒ **None** | |
| **11** | Stock or stock options | ☒ **None** | |
| **12** | Receipt of equipment, materials, drugs, medical writing, gifts or other services | ☒ **None** | |
| **13** | Other financial or non-financial interests | ☐ **None** | |
| | | F.M. is a full time employee at GeneDx, Inc. The authors declare no other conflicts of interest. | |

| | | Name all entities with whom you have this relationship or indicate none (add rows as needed) | Specifications/Comments (e.g., if payments were made to you or to your institution) |
|---|---|---|---|

**Please place an "X" next to the following statement to indicate your agreement:**

☒ I certify that I have answered every question and have not altered the wording of any of the questions on this form.
